# High COVID-19 transmission potential associated with re-opening universities can be mitigated with layered interventions

**DOI:** 10.1101/2020.09.10.20189696

**Authors:** Ellen Brooks-Pollock, Hannah Christensen, Adam Trickey, Gibran Hemani, Emily Nixon, Amy Thomas, Katy Turner, Adam Finn, Matt Hickman, Caroline Relton, Leon Danon

## Abstract

**Background:** Re-opening universities while controlling COVID-19 transmission poses unique challenges. UK universities typically host 20,000 to 40,000 undergraduate students, with the majority moving away from home to attend. In the absence of realistic mixing patterns, previous models suggest that outbreaks associated with universities re-opening are an eventuality.

**Methods:** We developed a stochastic transmission model based on realistic mixing patterns between students. We evaluated alternative mitigation interventions for a representative university.

**Results:** Our model predicts, for a set of plausible parameter values, that if asymptomatic cases are half as infectious as symptomatic cases then 5,760 (3,940 – 7,430) out of 28,000 students, 20% (14% – 26%), could be infected during the first term, with 950 (656 – 1,209) cases infectious on the last day of term. If asymptomatic cases are as infectious as symptomatic cases then three times as many cases could occur, with 94% (93% – 94%) of the student population getting infected during the first term. We predict that one third of infected students are likely to be in their first year, and first year students are the main drivers of transmission due to high numbers of contacts in communal residences. We find that reducing face-to-face teaching is likely to be the single most effective intervention, and this conclusion is robust to varying assumptions about asymptomatic transmission. Supplementing reduced face-to-face testing with COVID-secure interactions and reduced living circles could reduce the percentage of infected students by 75%. Mass testing of students would need to occur at least fortnightly, is not the most effective option considered, and comes at a cost of high numbers of students requiring self-isolation. When transmission is controlled in the student population, limiting imported infection from the community is important.

**Conclusions:** Priority should be given to understanding the role of asymptomatic transmission in the spread of COVID-19. Irrespective of assumptions about asymptomatic transmission, our findings suggest that additional outbreak control measures should be considered for the university setting. These might include reduced face-to-face teaching, management of student mixing and enhanced testing. Onward transmission to family members at the end of term is likely without interventions.

## Introduction

Despite the on-going COVID-19 epidemic, social distancing measures in many countries are beginning to be relaxed and universities across the world are due to start the new academic term from September 2020. In the UK, there are 2.3 million students, with up to 40,000 undergraduates at a single institution[1]. Universities are integral to many towns and cities in the UK: for example, in the 2011 census, a quarter of Oxford’s adult population was registered as a full-time student. Managing universities is a complex operation, and in the context of the COVID-19 epidemic, re-opening universities poses particular challenges for containing disease transmission.

Since June 2020, the UK has moved from a national containment strategy to localised containment of outbreaks, with the number of cases highly variable across the country. The imposition of lockdown in March 2020 led to a substantial reduction in travel and mobility, and local lockdowns have led to further reduced movement in some parts of the country. In the UK, re-opening universities is associated with a mass travel event. Around 80% of students leave home to attend University, moving an average 90 miles[2]. This synchronised event will increase population mixing at a national scale with the potential to spark outbreaks in new areas if not carefully managed. Once the university term starts there are more unique challenges facing universities. Students, in common with other 20-to-30-year olds, report high numbers of social contacts in their everyday lives[3]. Student accommodation frequently involves communal living, either in halls of residence that house several hundred students, or in all-student households renting in the private sector. Regular face-to-face teaching can involve several hundred students in a lecture theatre, and even without large lectures, tutorials and small group teaching involve close and prolonged contact between individuals.

The potentially high rate of transmission within a university setting is unlikely to translate to high morbidity among students. There is a marked age disparity in severe COVID-19 cases, with younger people less likely to exhibit typical symptoms or suffer severe outcomes[4]. In the UK, less than 0.2% of COVID-19-related deaths are in persons under 30. Students are typically young adults in their early twenties. Nevertheless, it appears likely that young adults are susceptible to infection and infectious to others. Hence there is a risk of asymptomatic transmission within the student population, posing a risk to vulnerable students, people outside the university setting and family members when students return home.

A number of studies have investigated the challenges inherent in reopening of universities amidst the COVID-19 pandemic[5,6]. Existing models have focused on isolated campus universities in the US, rather than civic universities that are common in the UK and elsewhere[6], and the majority have not had access to realistic mixing patterns within the university setting, which drive transmission. In this paper we combined analysis of social contact data with a data-driven mathematical modelling approach to investigate the impact of re-opening a UK university on COVID-19 transmission. We characterise patterns of disease transmission and investigate potential mitigating effects of interventions. These results are used to synthesise guidance on measures that universities might wish to consider for effective outbreak control once students arrive or return for the forthcoming academic year.

## Methods

### Data sources

We used two datasets for our analysis: a re-analysis of the Social Contact Survey (SCS)[3,7] and a data extract from the University of Bristol (UoB).

### Social Contact Survey data

The SCS was a paper-based and online survey of 5,388 participants in Great Britain conducted in 2010[3,7]. We have previously used these data to estimate the reproduction number for COVID-19[8] [ref]. The SCS included 363 participants whose listed occupation included “STUDENT”. We extracted these participants to summarise their contacts by context (home, university, leisure/other, travel) and to estimate the potential COVID-19 reproduction number in students. We used a Student’s t-test to determine the level of evidence for the observed differences in numbers of contacts between students and the general population.

We used the SCS to estimate the contact rate between students by year and school. For a student in group *i*, we took the number of study contacts as 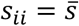 and *S_ij_* = 0, where 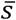 is the mean number of university-associated contacts reported by students in the SCS. For non-study contacts, we took *r_ij_* = 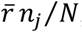, where 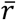, is the mean number of other/leisure contacts reported by students in the SCS and *n_j_*/*N* is the proportion of students in group *j*.

### University data

We were provided with an anonymised extract of student data for a university relating to the

2019/2020 academic year. The study complied with the University data protection policy for research studies (http://www.bristol.ac.uk/media-library/sites/secretary/documents/information-governance/data-protection-policy.pdf).

The data contained age, primary faculty affiliation (7 faculties), primary school affiliation (28 schools), year of study (6 undergraduate years, taught postgraduates and research postgraduates), term-time residence, home region (if in the UK), and country of origin for students registered in 2019/2020.

We used the university data to estimate the household contact rate between students by year and school. We estimated the number of household contacts from the student data, taking postcode as a proxy for household. The average number of students in school/year group *j* sharing accommodation with a student in group *i* is calculated as:

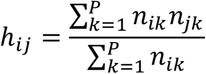

where *n_ik_* is the number of students in school/year *i* living at unique postcode *k* and *P* is the number of unique postcodes.

In UoB, students in university residences will be assigned to a living circle, which is a group of students that have higher rates of contact. We take the baseline living circle size as 24 students and investigate the impact of smaller living circles. Where the number of students at a single postcode exceeded the living circle size, we create subunits within the postcode that are the size of the living circle. Each living circle contains a random sample of students at that address. See supplementary figure S1 for a pictorial explanation of how the data are processed.

### Modelling framework

We use a stochastic compartmental model to simulate transmission dynamics in the student population at UoB. We assumed that COVID-19 could be captured by seven infection states: susceptible to infection (S), latently infected (E), asymptomatic and infectious (A), pre-symptomatic and infectious (P), symptomatic and infectious (I), self-isolating (Q), hospitalised (H) and recovered and immune (R). The total number of students is given by *N*. The flow between compartments is depicted in figure 1 and given by the equations below.

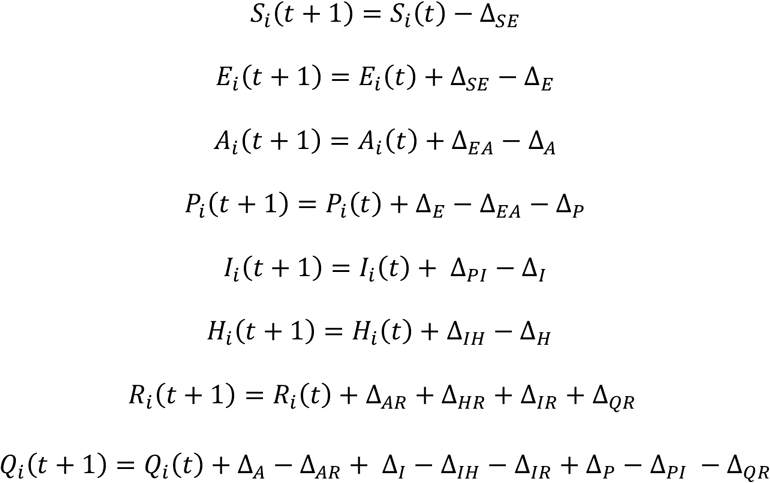

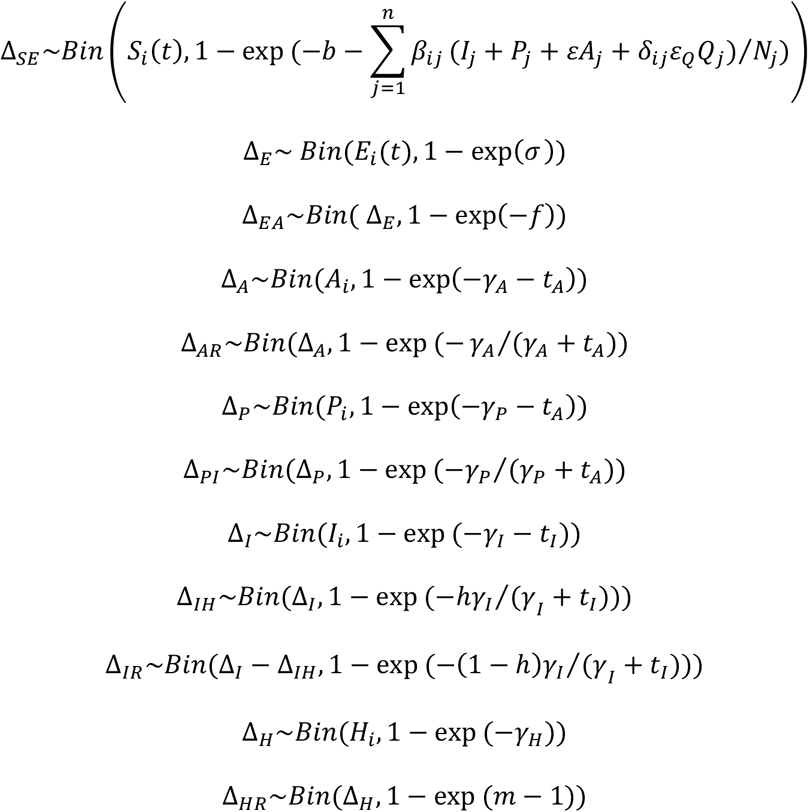

**Figure 1:**
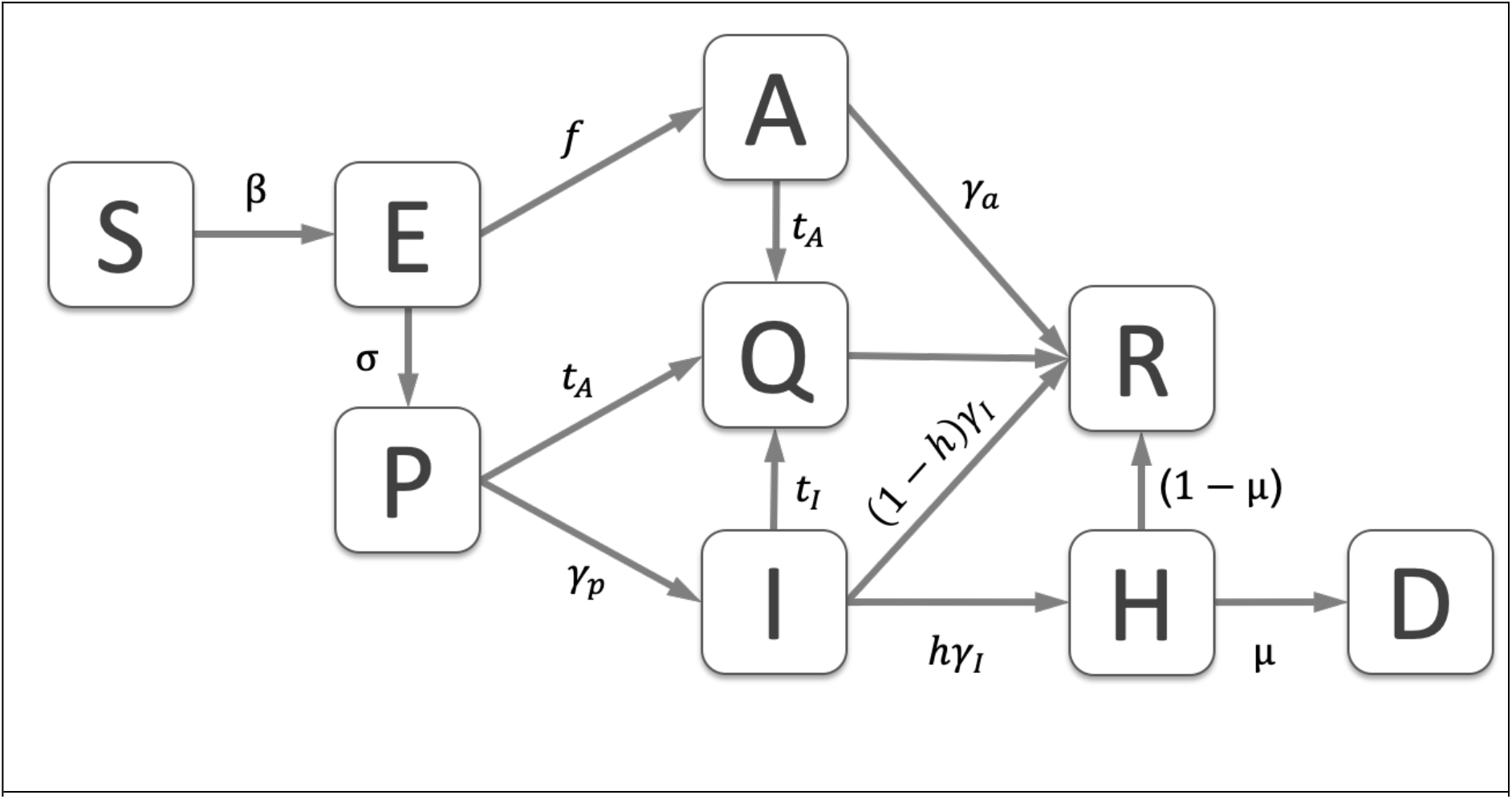
a) Model flow diagram with infection states and rates between them.

### Model parameters

The student population was divided into 161 groups representing school and year of study. The proportion of students in each group and the mixing between groups was taken from the mixing matrix in figure 2. As 92% of the student population is under 30 years of age, we expect a high proportion of cases to be asymptomatic[9] (*f* = 0.75), a low hospitalisation rate[10] (*h* = 0.002) and a low mortality rate of hospitalised cases (*μ* = 0.038)[10]. We assume that only symptomatic cases can be hospitalised, and only hospitalised cases can die.

**Figure 2:**
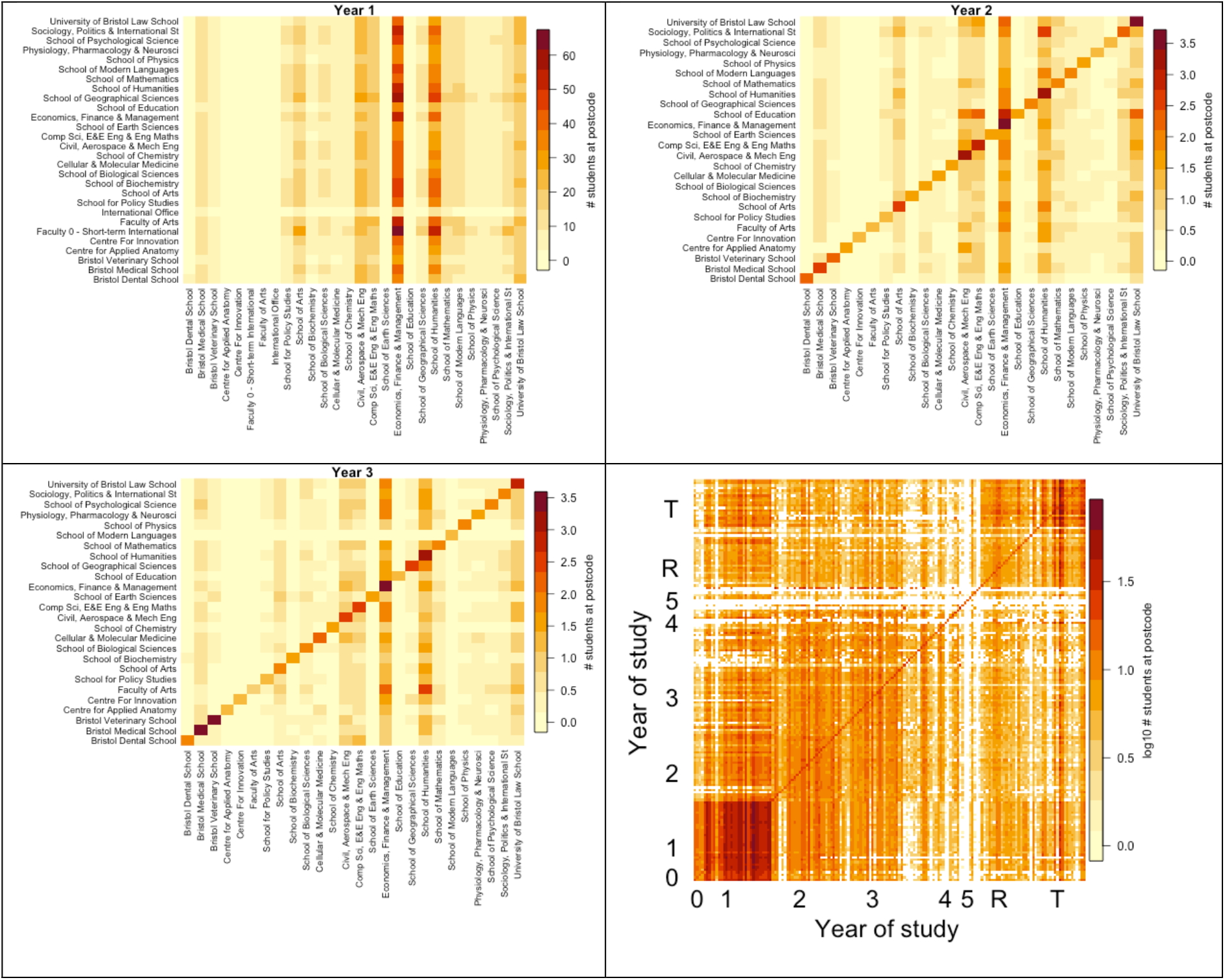
The average number of students sharing accommodation by school (a) year 1; (b) year 2; (c) year 3; (d) for all years and schools.

For symptomatic cases, we assume an average incubation period, during which cases are assumed not to be infectious and cannot be detected by the test, of 〈1/*σ*〉 = 3 days[11], 3 days[11], after which they become infectious but pre-symptomatic for a period of 〈1/*γ*〉 = 2 days, when cases can be detected with a test. The infectious period is taken as 〈1/*γ*〉 = 3 days[11], although there is uncertainty around these values. We take the number of days in hospital as 〈1/*γ_H_*〉 =7 days[12]. Symptomatic individuals are tested and moved to self-isolation at a rate *t_I_*.

Asymptomatic cases are infectious for 〈1/*γ*〉 = 5 days, so that their average infectious period equals the infectious period for symptomatic cases. Asymptomatic cases are tested and moved to self-isolation at rate *t_A_* where they remain for an average of 14 days. Individuals in self-isolation contribute to the force-of-infection within their subgroup only at a reduced rate *ε_Q_* = 0.5.

The infectiousness of asymptomatic cases relative to symptomatic cases is represented by the parameter *ε*. It is now generally accepted that asymptomatic transmission can and does occur, however much of it appears to be pre-symptomatic transmission, i.e. in the days before symptom onset [13]. A modelling study estimated that transmission due to truly asymptomatic cases (i.e. cases that never go on to develop symptoms) was limited, with pre-symptomatic and symptomatic transmission contributing the remainder in approximately equal proportions[14]. We take a baseline value for the relative infectiousness of asymptomatic cases, *ε* of 0.5 and consider values between 0 and 1. We assume that pre-symptomatic and symptomatic cases are equally infectious[15].

We assume the transmission rate between group *i* and group *j*, *β_ij_*. is proportional to the contact rate *c_ij_*, where *c_ij_* is the average number of contacts in group *j* made by a person in group *i*. We assume that contacts were either household contacts (*h_ij_*), study contacts (*s_ij_*) or random contacts (*r_ij_*), so each entry in the contact matrix is given by *c_ij_* = *h_ij_* + *s_ij_* + *r_ij_*. In order to translate the contact matrix into the transmission matrix, we calculate a constant *k* such that the maximum eigenvalue of the transmission matrix B = {*β_ij_*} = {*kc_ij_*} equals the reproduction number[16].

To estimate the reproduction number in the student population, we took a population-wide reproduction number of *R*_0_ = 2.7. In our framework, if a symptomatic case generates *R_s_* secondary cases, then an asymptomatic case will generate *R_A_* = *εR_S_* secondary cases. With *R*_0_ = *R_S_* + *R_A_*, *R_S_* = *R*_0_/(*f* + (1 − *f*)*ε*). If cases without symptoms are 50% as infectious as cases with symptoms (*ε* = 0.5), and a fraction *f* = 0.6 of the general population has symptoms when infected, then in a university setting when a lower proportion of cases have symptoms (*f* = 0.25) but have on average 10% more contacts than an average person, we would expect a reproduction number within university of *R_U_* = 2.7. If *ε* = 0.1 then *R_U_* = 1.7; if *ε* = 1 then *R_U_* = 3.4 (see SI, section 2, figure S2).

### Initial conditions and model implementation

We estimated the number of infected students at the start of term using home location and incidence in home location as of July 2020. For each scenario, we ran the model 100 times using a different random seed. The model was simulated for one year to illustrate the full range of dynamics, and we consider the state of the outbreak after 84 days, which is the number of days between the start of the September term and the winter holidays at the end of the first term. The model code is available at https://github.com/ellen-is/unimodel.

### Sensitivity analysis

We explored the inherent variability of the model by running the model with baseline parameters for 100 realisations, and then running 100 more realisations of the model varying the baseline parameters by +/−10%.

The impact of the infectiousness of asymptomatic cases was explored for values of *ɛ* between 0 (asymptomatic cases not infectious) and 1 (asymptomatic cases as infectious as symptomatic cases), which corresponds to reproduction numbers ranging from 1.7 to 3.4 (see SI section 2).

### Control options

We assumed that the university would be operating within Public Health England (PHE) guidelines, i.e. that symptomatic cases should be tested and self-isolate within 48 hours. Contact tracing is difficult to implement explicitly in the compartmental model framework, but the mechanism of action can be captured by a lower within-group transmission rate. We focussed on interventions that could be implemented on top of wider control measures. We considered the following interventions (see table 1 for a summary):

- **Baseline** conditions are “business as usual” behaviour within universities with PHE guidelines. Symptomatic cases are tested are moved into self-isolation after an average of 48 hours if test positive. No additional testing for people with no symptoms. Students are assumed to be in living circles that comprise of a maximum of 24 individuals to reflect existing UoB arrangements.
- **COVID security** is modelled by reducing the transmission probability associated with non-residence contacts by 25% and 50% to capture the impact of face covering use and social distancing.
- **Reduced face-to-face teaching** is captured by reducing the number of face-to-face teaching contacts from 20 students to 15 and then 5 students.
- **Reduced living circles** reflects reducing the number of students sharing facilities within accommodation. In the baseline scenario, we assumed that students were in contact with other students living in the same accommodation, forming household groups up to a maximum of 24 individuals. For accommodation with more than 24 residents, we divided the accommodation population up into subunit “living circles” of 24 students. To explore the impact of living circle size, we reduced the maximum living circle size from 24 to 20 and then 14 persons.
- **Reactive mass testing**: We simulate scenarios in which all students are tested the presence of current infection if the number of test-positive cases in a given week is greater than the previous week. We move test positive cases into self-isolation after an average of 2 days. Test all students within 2 to 28 days if the number of test-positive cases increases from one week to the next. Additional testing is continued until the number of test-positive cases in a given week is less than the previous week.
- **Multiple, layered interventions:** We investigated the impact of each of the above interventions in isolation and then applied sequentially: 25% reduction in transmission due to COVID security, followed by a reduction in face-to-face teaching to 5 study contacts, followed by a reduction in living circles to 24 individuals, and reactive mass testing every 2 days if the infection rate on campus should rise, and finally a reduction in importation rates from outside the university population.

**Table 1:** Intervention scenarios for controlling transmission within university settings.

| Scenario | Transmission probability per contact household/other | # random contacts | # within-course contacts | Max living circle size | % transmission reduction due to self-isolation within/between groups | Asymptomatic testing |
| --- | --- | --- | --- | --- | --- | --- |
| Baseline | 0.05<sup>§</sup> | 4* | 20* | 24 | 50% / 100% | None |
| COVID security | 0.05<sup>§</sup> / 0.04 or 0.025 | 4* | 20* | 24 | 50% / 100% | None |
| Reduced face-to-face teaching | 0.05<sup>§</sup> / 0.05<sup>§</sup> | 4* | 15 or 5 | 24 | 50% / 100% | None |
| Reduced living circle size | 0.05<sup>§</sup> / 0.05<sup>§</sup> | 4* | 20* | 20 or 14 | 50% / 100% | None |
| Improved self-isolation | 0.05<sup>§</sup> / 0.05<sup>§</sup> | 4* | 20* | 24 | 100% / 100% | None |
| Reactive mass testing | 0.05<sup>§</sup> / 0.05<sup>§</sup> | 4* | 20* | 24 | 50% / 100% | Every 2 or 7 days when rates are increasing |
| Multiple | 0.05<sup>§</sup> / 0.04 | 4* | 5 | 14 | 50% / 100% | Every 2 days when rates are increasing |

\*Estimated from the Social Contact Survey, <sup>§</sup>calculated such that $R=2.7$ .

For each model realisation, we calculated the doubling time during the exponential growth phase as ln (2)/*r*, where *r* is the exponential growth rate in the number of infected individuals, the incident number of symptomatic and asymptomatic cases at the end of the first term (day 84 of the model), the time the outbreak turns over, the number of students in self-isolation and ratio of asymptomatic to symptomatic cases.

We ranked the interventions when implemented without additional measures by mean number of symptomatic cases at the end of the first term calculated from 100 realisations of the model for a given set of parameters, and repeated this ranking for values of *ε* between 0 (asymptomatic cases not infectious) and 1 (asymptomatic cases as infectious as symptomatic cases).

## Results

### Contact patterns and estimated reproduction number in university students

The Social Contact Survey included 363 participants whose listed occupation included “STUDENT”. Students reported more home contacts than other participants (3.5 versus 2.3, p-value < 0.001). However, although students reported more contacts than other participants on average, there was no evidence of a systematic difference (29.9 versus 26.8, p-value 0.40). The majority (82% 95% CI: 79% to 86%) of students’ social contacts are either home or associated with university. On average, students reported 20.0 (95% CI: 14.1, 28.8) university contacts, and 4.3 (95% CI: 2.7, 6.5) other/leisure contacts.

Although students do not have more contacts than the general population, 18 to 24-year olds do have more contacts than the wider population. Taken in combination with the contact duration, the individual reproduction number for this age group is higher than for other individuals.

### Heterogeneous mixing rates within a university

To capture student contact patterns within a university, we used comprehensive anonymised student accommodation data for the academic year 2019/2020 from UoB. The data included 20,819 registered undergraduates and 8,501 registered postgraduates divided into 6 faculties and 28 schools and 2,862 unique postcodes (see supplementary table 1 for number of students by year of study and faculty). Most students (92%) are under 30 years of age and the largest school is the School of Economics, Finance and Management with 3,674 students.

We used the student data to create synthetic contact matrices for mixing between year groups and schools. From postcodes we generated between school contact matrices for each year of study, and for all years (figure 2). Halls of residence dominate the first-year contact matrix, with mixing across all schools and no clear assortative mixing (figure 2a).

In years 2 and 3 the average household size decreases substantially and there is increased assortativit mixing between schools, indicating that students are more likely to share accommodation with someone from their own school by choice (figures 2b & 2c).

The university-wide contact matrix consists of 161 groups of students categorised by 28 schools and nine year-groups (0, 1, 2, 3, 4, 5, 6, PGT, PGR) (figure 2d). The higher levels of mixing between first years is evident in the lower left-hand corner and the assortative mixing by year and school is shown by the diagonal. There are fewer inter-year household contacts and more intra-university mixing between taught postgraduates than for research postgraduates.

### Transmission dynamics in the student population

We investigated the dynamics of an epidemic with limited mitigation in the student population with plausible COVID-19 parameter values and assuming that symptomatic cases are tested and self-isolate within 48 hours. Because of the population structure, the stochasticity and relatively small numbers involved, there is large intrinsic variability between simulations with identical parameter values; we report the mean, minimum and maximum from 100 simulations.

Using plausible parameters (asymptomatic cases half as infectious as symptomatic cases and a reproduction number of *R_U_* = 2.7), and without interventions or holidays, we predict a university-wide outbreak with an early growth rate of 0.07 (0.03-0.10), which is equivalent to a doubling time of 9 days (7-24 days) (figure 3a). Based on the timescales of COVID-19 with baseline parameters, we expect that it would take around 4 months for the outbreak to peak, assuming no winter break.

**Figure 3:**
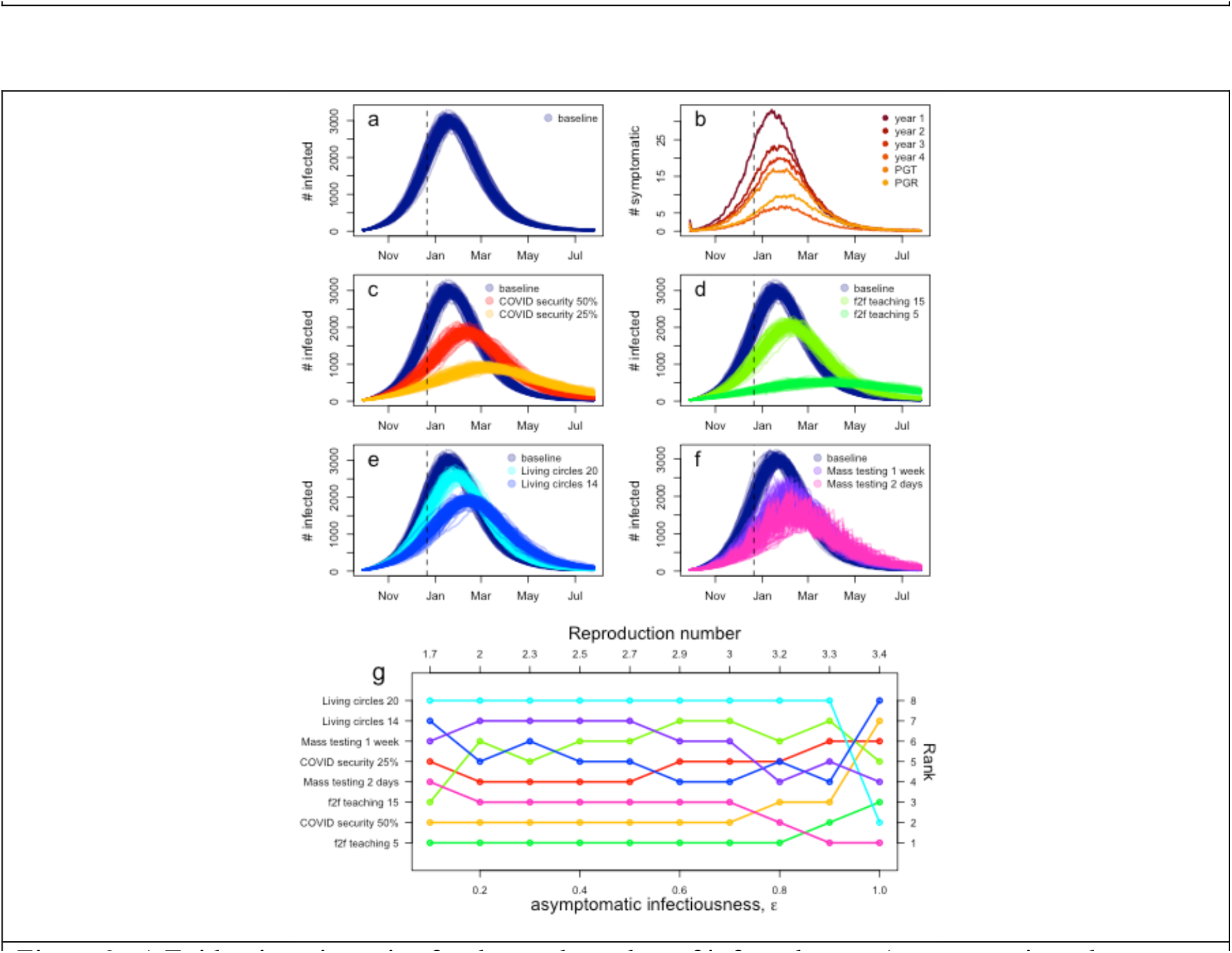
a) Epidemic trajectories for the total number of infected cases (symptomatic and asymptomatic cases) the baseline model from 100 realisations with best estimate parameters; b) Mean number of symptomatic cases by year group from 100 realisations; c) Epidemic trajectories when COVID security measures reduce transmission by 50% and 25%; d) Epidemic trajectories when face-to-face teaching is limited to 15 and 5 persons; e) Epidemic trajectories for reduced living circles to 20 and 14 persons; e) Epidemic trajectories when reactive mass testing is implemented every week and every 2 days. Dotted vertical lines denote the end of the first term. g) Ranking of interventions by mean number of symptomatic cases at the end of the first term from 100 realisations for increasing values of asymptomatic infectiousness, and therefore also increasing values of the reproduction number. The colours correspond to the colours of the epidemic trajectories above.

First year students drive the early part of the outbreak and experience the highest burden of infection, followed by second and third years and taught postgraduate students (figure 3b). Students in year 4 and above and research postgraduates have the lowest infection rates.

By the end of the first term, 5,760 (3,940 – 7,430) out of 28,000 students, or 20% (14% – 26%), have been infected with 73 (47 – 100) symptomatic cases and 880 (610 – 1,110) asymptomatic cases infectious on the last day of term. Without additional control measures, 74% (72% – 75%) of students would be infected by the end of the academic year. The low rate of symptoms and low morbidity rate results in a median of zero deaths in the student population.

The relative infectiousness of asymptomatic cases is central for determining the scale of a university-based outbreak. In our framework, asymptomatic cases are either less or as infectious as symptomatic cases, however because asymptomatic cases do not self-isolate without a test, for higher values of relative infectiousness, *ε*, asymptomatic cases produce on average more secondary cases than symptomatic cases (see SI figure S2). For lower values of *ε* university-focussed outbreaks are largely driven by the forcing from outside the university. For intermediate values, outbreaks peak after the first term. For high values, outbreaks peak before the end of the first term (figure S4)

As a comparison to the baseline case, if asymptomatic cases are as infectious as symptomatic cases (*R_U_* = 3.4) then we expect an early growth rate of 0.12 (0.10 – 0.14) and a doubling time of 5.8 days (5 – 7 days). Without additional control measures, 96% (95% – 97%) of the student population would be infected by the end of the academic year. If asymptomatic cases are 30% as infectious as symptomatic cases (*R_U_* = 2.25) then we expect an early growth rate of 0.06 (0.04 – 0.09) and a doubling time of 12 days (8 – 17 days). Without additional control measures, 28% (20% – 35%) of the student population would be infected by the end of the academic year. The epidemic profiles for the full range of potential scenarios for asymptomatic infectiousness, which corresponds to reproduction numbers from 1.7 to 3.4 are shown in supplementary figure S4.

### University-based interventions that mitigate transmission

We investigated multiple interventions that reduced the infection burden in the student population (figures 3c – f). The impact of implementing each intervention was explored in isolation and in combination with other measures. When layering interventions, we implemented lower cost interventions first, such as creating COVID-secure interactions with face coverings and social/physical distancing, and reserved mass testing of non-symptomatic students as a more resource-intensive intervention.

For realistic values of COVID security and *R_U_* = 2.7, we find that reducing the transmission probability with COVID secure interactions has the potential to reduce, but not completely eliminate, the size of outbreaks (figure 3c). We estimate that by reducing transmission for non-household contacts by 25% the early doubling time is increased slightly to 11 (8 – 17) days. The percentage of students infected by the end of the first term is 10% (7% – 13%) and the number of symptomatic and asymptomatic students infectious on the last day of term is decreased to 35 (15 – 53) and 410 (270 – 550) asymptomatic cases. Reducing transmission for non-household contacts by 50% increases the doubling time to 12 (9 – 17) days and further reduces the number of infectious students on the last day of term 15 (10 – 22) symptomatic cases and 200 (160 – 260) asymptomatic cases.

Reducing the number of interactions made during face-to-face teaching from 20 to 15 other students increases the early doubling time to 10 (7 – 22) days and reduces the number of infected students at the end of the first term to 39 (19 – 60) symptomatic cases and 460 (320 – 620) (figure 3d). Reducing the number of face-to-face contacts from 20 to 5 other students was the single most impactful intervention investigated in terms of the number of students infected by the end of the first term and the number of infectious students on the last day of term, increasing doubling time to 14 (9 – 28) days, including scenarios in which the number of cases in the student population was driven to zero (figure 3d). The number of infected students at the end of the first term was 11 (4 – 20) symptomatic cases and 130 (80 – 200) asymptomatic cases.

Implemented without other measures, reducing the size of living circles (defined as the number of students that share bathroom/kitchen facilities) from 24 to 20 or 14 students was overall the least effective intervention investigated (figure 3e, supplementary information Table 2). However, when implemented on top of COVID secure interactions and reductions in face-to-face teaching, reducing living circles to 14 individuals does reduce the total percentage of students that are infected by the end of the first term by 25%.

Mass testing all students regardless of symptoms was effective at reducing the total number of infections and the initial rate of epidemic growth rate, but reactive testing was required for the whole year (figure 3f). Compared to other interventions, mass testing was generally more effective for higher values of the reproduction number and resulted in the third lowest number of infected students by the end of the first term. However, for lower values of asymptomatic infectiousness, and hence lower values of the reproduction number, reducing face-to-face teaching, implementing COVID security and reduced living circles was more effective than testing all students (figure 3g).

Testing all students primarily reduced the number of students with asymptomatic and presymptomatic infections, reducing the ratio of asymptomatic to symptomatic cases to 9:1 (8:1 – 10:1). However, the reduction in infection from mass testing comes at a substantial cost in terms of the number of students self-isolating: under 2 day testing, at the height of the outbreak 1,300 (860-1,500) students (4.5%, 3% – 5%) were self-isolating compared to 520 (470 – 560) students (1.9%, 1.7% – 2.0%) in the baseline scenario.

Testing all students monthly had a minimal impact compared to not testing at all, reducing the average percentage of students infected during the outbreak by 1.3%. Increasing testing frequency to fortnightly, weekly or every 3 or 2 days was beneficial, and this was robust to parameter choice (supplementary figure S5).

We found that implementing multiple, layered interventions was able to effectively control transmission in the student population (figure 4a-c). The remaining cases in students were largely due to importation of infection from outside the university setting: reducing the background rate of infection demonstrates that if imported infections could be managed then the number of infected students could be very low.

**Figure 4:**
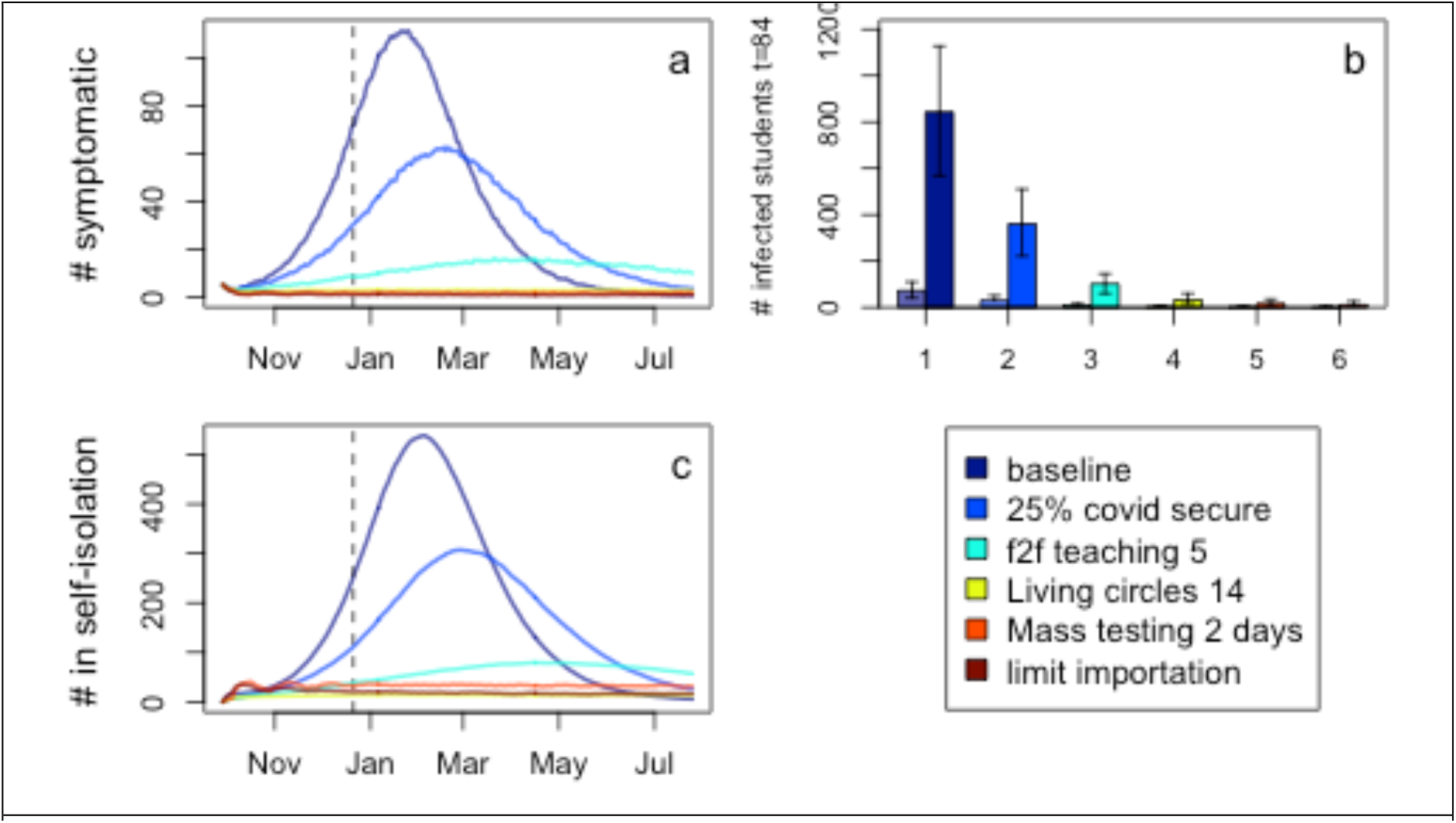
Impact of implementing multiple interventions sequentially. a) Number of symptomatic cases; b) Number of infected (symptomatic and asymptomatic) students at the end of the first term (day 84); c) Number of students that are self-isolating.

## Discussion

Our results suggest that, under normal circumstances, COVID-19 would spread readily in a university setting. Our data-driven approach reveals natural heterogeneities in student mixing patterns that can be exploited to enhance disease control. We find that controlling transmission is possible with combinations of social distancing, online teaching, self-isolation, and potentially mass testing of students without symptoms.

Our findings highlight the importance of monitoring first year students and halls of residence in particular. In our analysis, first year students experienced the highest rates of infection and dominate the early part of the outbreak due to the high levels of mixing in halls of residence. Halls of residence have been identified as a risk factor for the transmission of close contact infections including meningitis[17], mumps[18], norovirus[19], respiratory illnesses[20] and gastroenteritis[21]. In practice, students in larger residences are allocated into shared flats or living circles, potentially limiting widespread transmission. Maintaining social distancing between living circles within residences is paramount for maintaining COVID-19 control.

Lessons about infection control in universities can be learnt from other diseases. Mass vaccination used for meningitis, mumps and rubella outbreaks is not an option for COVID-19 at present. During a mumps outbreak in a university hall of residence, Kay et al (2011) reported difficulty in identifying higher risk students[18]. Due to the high number of contacts and of students’ contact networks inhabit, universities may wish to consider how they might facilitate the collation of data to expedite the contact tracing process. Embedding positive health behaviours like hand washing and using face coverings will also contribute to minimising transmission opportunities. A randomised control trial of hand washing in university residences found that installing alcohol hand sanitizer in every room, bathroom and dining hall reduced respiratory illness in students by 20%[20].

Previous modelling work, based on universities in the United States, has focussed on the necessity of regularly testing all students[5]. While our findings are consistent that frequent testing is necessary if used in isolation, our modelling approach demonstrates that other interventions are viable. This is partly due to our result that suggests that the reproduction number would be lower than previous studies have assumed due to the high proportion of asymptomatic cases. Furthermore, as previous studies have discussed[5], when prevalence is low the false positive rate can exceed the true positive rate leading to unnecessary isolation of negative cases. We tried to mitigate false positive cases by implementing reactive mass testing once incidence increased; an alternative approach would be to use a second confirmatory test. Antibody testing could also play in role in determining prior infection and infection rates in student populations.

Our work uses a similar compartmental modelling approach to the handful of models that have been developed for COVID-19 transmission in universities in the USA. A drawback of this approach is that individual behaviour is not readily captured; in particular, contact tracing and isolation of contacts or living circles is difficult to include in detail. We capture some heterogeneity using household and faculty mixing data, and a stochastic model was necessary due to the potentially small number of students in each subgroup. Nevertheless, a network modelling approach would be more appropriate for studying superspreading events and individual-level variation.

Furthermore, while we had detailed data pertaining to the university student population, we had limited data on contact with the location population and we did not include university staff explicitly in the model. Given the age distribution of students, and the high likelihood of asymptomatic infection, staff and surrounding communities are likely to experience higher levels of morbidity than the students themselves. Although by-and-large students fraternise with students, they do pose some risk to more vulnerable groups within the university such as staff with co-morbidities, or to their local community. Safeguarding all is a high priority.

The aim of this work was to characterise potential COVID-19 transmission patterns in a university setting and identify strategies that may prove more likely to control transmission. In the absence of university outbreak data, we used COVID-19 transmission parameters estimated from other settings. Once the university year starts, and should there be an outbreak, this type of modelling should be used to estimate parameters in real time and provide a more accurate tool for guiding interventions.

## Data Availability

Contact the corresponding author regarding the data used for modelling.

## Acknowledgements

We thank TJ McKinley for technical advice and comments, Gabor Soter for his contribution and the University of Bristol for providing the data.

## References

1. Universities UK. Patterns and Trends in UK Higher Education. Focus Univ UK. 2018. Available: http://www.universitiesuk.ac.uk/highereducation/Documents/2013/PatternsAndTrendsinUKHigherEducation2013.pdf

2. Higher Education Statistics Agency. Full-time and sandwich students by term-time accommodation 2014/15 to 2018/19. 2019. Available: https://www.hesa.ac.uk/data-andanalysis/students/chart-4

3. Danon L, Read JM, House TA, Vernon MC, Keeling MJ. Social encounter networks: characterizing Great Britain. Proc Biol Sci. 2013; 280: 20131037. doi: 10.1098/rspb.2013.1037

4. Sinha IP, Harwood R, Semple MG, Hawcutt DB, Thursfield R, Narayan O, et al. COVID-19 infection in children. Lancet Respir Med. 2020; 8: 446–447. doi: 10.1016/S2213-2600(20)30152-1

5. Paltiel AD, Zheng A, Walensky RP. Assessment of SARS-CoV-2 Screening Strategies to Permit the Safe Reopening of College Campuses in the United States. JAMA Netw open. 2020; 3: e2016818. doi: 10.1001/jamanetworkopen.2020.16818

6. Christensen H, Turner K, Trickey A, Booton RD, Hemani G, Nixon E, et al. COVID-19 transmission in a university setting: a rapid review of modelling studies. medRxiv. 2020; 2020.09.07.20189688. doi: 10.1101/2020.09.07.20189688

7. Danon L, House TA, Read JM, Keeling MJ. Social encounter networks: collective properties and disease transmission. J R Soc Interface. 2012; 9: 2826–2833. doi: 10.1098/rsif.2012.0357

8. Brooks-Pollock E, Read JM, McLean A, Keeling MJ, Danon L. Using social contact data to predict and compare the impact of social distancing policies with implications for school reopening. medRxiv. [cited 8 Sep 2020]. doi: 10.1101/2020.07.25.20156471

9. Viner RM, Mytton OT, Bonell C, Melendez-Torres GJ, Ward JL, Hudson L, et al. Susceptibility to and transmission of COVID-19 amongst children and adolescents compared with adults: a systematic review and meta-analysis. medRxiv. 2020; 2020.05.20.20108126. doi: 10.1101/2020.05.20.20108126

10. Dorigatti I, Okell L, Cori A, Imai N, Baguelin M, Bhatia S, et al. Report 4: Severity of 2019-novel coronavirus (nCoV). 2020.

11. Li Q, Guan X, Wu P, Wang X, Zhou L, Tong Y, et al. Early transmission dynamics in Wuhan, China, of novel coronavirus-infected pneumonia. New England Journal of Medicine. 2020. pp. 1199–1207. doi: 10.1056/NEJMoa2001316

12. Docherty AB, Harrison EM, Green CA, Hardwick HE, Pius R, Norman L, et al. Features of 20 133 UK patients in hospital with covid-19 using the ISARIC WHO Clinical Characterisation Protocol: Prospective observational cohort study. BMJ. 2020; 369. doi: 10.1136/bmj.m1985

13. Huff H V, Singh A. Asymptomatic Transmission During the Coronavirus Disease 2019 Pandemic and Implications for Public Health Strategies. Clin Infect Dis. 2020 [cited 8 Sep 2020]. doi: 10.1093/cid/ciaa654

14. Ferretti L, Wymant C, Kendall M, Zhao L, Nurtay A, Abeler-Dörner L, et al. Quantifying SARS-CoV-2 transmission suggests epidemic control with digital contact tracing. Science (80-). 2020; eabb6936. doi: 10.1126/science.abb6936

15. Lipman M, White J. Collaborative tuberculosis strategy for England. Bmj. 2015; 350: h810–h810. doi: 10.1136/bmj.h810

16. Hens N, Goeyvaerts N, Aerts M, Shkedy Z, Van Damme P, Beutels P. Mining social mixing patterns for infectious disease models based on a two-day population survey in Belgium. BMC Infect Dis. 2009; 9: 5.

17. Gilmore A, Jones G, Barker M, Soltanpoor N, Stuart JM. Meningococcal disease at the University of Southampton: Outbreak investigation. Epidemiol Infect. 1999; 123: 185–192. doi: 10.1017/S0950268899002794

18. Kay D, Roche M, Atkinson J, Lamden K, Vivancos R. Mumps outbreaks in four universities in the North West of England: Prevention, detection and response. Vaccine. 2011; 29: 3883–3887. doi: 10.1016/j.vaccine.2011.03.037

19. Bhatta MR, Marsh Z, Newman KL, Rebolledo PA, Huey M, Hall AJ, et al. Norovirus outbreaks on college and university campuses. J Am Coll Heal. 2019 [cited 17 Aug 2020]. doi: 10.1080/07448481.2019.1594826

20. White C, Kolble R, Carlson R, Lipson N, Dolan M, Ali Y, et al. The effect of hand hygiene on illness rate among students in university residence halls. Am J Infect Control. 2003; 31: 364–370. doi: 10.1016/S0196-6553(03)00041-5

21. Moe CL, Christmas WA, Echols LJ, Miller SE. Outbreaks of acute gastroenteritis associated with norwalk-like viruses in campus settings. J Am Coll Health Assoc. 2001; 50: 57–66. doi: 10.1080/07448480109596008

